# ACE-inhibitors and Angiotensin-2 Receptor Blockers are not associated with severe SARS-COVID19 infection in a multi-site UK acute Hospital Trust

**DOI:** 10.1101/2020.04.07.20056788

**Authors:** Daniel M Bean, Zeljko Kraljevic, Thomas Searle, Rebecca Bendayan, Kevin O’Gallagher, Andrew Pickles, Amos Folarin, Lukasz Roguski, Kawsar Noor, Anthony Shek, Rosita Zakeri, Ajay M Shah, James TH Teo, Richard JB Dobson

**Author notes:** joint author (Ajay M Shah, James TH Teo, Richard JB Dobson). joint corresponding authors: Prof Richard Dobson, Dept of Biostatistics and Health Informatics, Institute of Psychiatry, Psychology and Neuroscience, King’s College London, 6 De Crespigny Park, London SE5 8AF, UK. Or: Dr Dan Bean; Dr James Teo; Prof Ajay Shah.

## Abstract

**Aims:** The SARS-Cov2 virus binds to the ACE2 receptor for cell entry. It has been suggested that ACE-inhibitors (ACEi) and Angiotensin-2 Blockers (ARB), which are commonly used in patients with hypertension or diabetes and may raise ACE2 levels, could increase the risk of severe COVID19 infection.

**Methods and Results:** We evaluated this hypothesis in a consecutive cohort of 1200 acute inpatients with COVID19 at two hospitals with a multi-ethnic catchment population in London (UK). The mean age was 68±17 years (57% male) and 74% of patients had at least 1 comorbidity. 415 patients (34.6%) reached the primary endpoint of death or transfer to a critical care unit for organ support within 21-days of symptom onset. 399 patients (33.3 %) were taking ACEi or ARB. Patients on ACEi/ARB were significantly older and had more comorbidities. The odds ratio (OR) for the primary endpoint in patients on ACEi and ARB, after adjustment for age, sex and co-morbidities, was 0.63 (CI 0.47-0.84, p<0.01).

**Conclusions:** There was no evidence for increased severity of COVID19 disease in hospitalised patients on chronic treatment with ACEi or ARB. A trend towards a beneficial effect of ACEi/ARB requires further evaluation in larger meta-analyses and randomised clinical trials.

## Introduction

The SARS-Cov2 pandemic is a major medical and socioeconomic challenge with at least 3 million confirmed cases to date. Data on the clinical characteristics of patients who require hospital admission for COVID19 disease from China, Italy and the US consistently show that patients with cardiovascular comorbidities are over-represented and may have an increased risk of severe COVID19 disease.^1-3^ The reasons underlying the increased incidence of severe COVID19 infection in those with comorbidities such as hypertension, diabetes and other cardiovascular conditions are unknown.

The SARS-Cov2 virus requires the binding of its viral surface spike protein to the ACE2 receptor expressed on epithelial cells in order to be internalised and then undergo replication.^4^ Previous studies suggest that the expression of ACE2 may be increased by chronic treatment with ACEi or ARB.^5^ As such, it has been hypothesized that treatment with ACEi or ARB could increase the likelihood of SARS-Cov2 binding and entry into epithelial or other cells.^6^ Furthermore, it is hypothesised that such a mechanism could account for the increased incidence of severe COVID19 infection among patients with cardiovascular comorbidities, who are frequently treated with ACEi/ARB.^6^ Whether or not treatment with ACEi/ARB increases the risk of severe COVID19 disease is a very important question in view of the large numbers of patients potentially on these drugs, especially in western countries with older populations. The issue is controversial because ACEi/ARB may potentially be beneficial in severe lung injury by reducing activation of the renin angiotensin system (RAS).^7,8^ Furthermore, increased levels of ACE2 itself have been shown to be protective during severe lung injury.^9,10^ The potential effect of ACEi and ARB during infection with SARS-CoV-2 therefore requires urgent clarification.

We tested for association between treatment with ACEi/ARB and disease severity in a consecutive series of 1200 patients with COVID19 disease admitted to two UK hospitals, King’s College Hospital and Princess Royal University Hospital, that have been at the epicentre of the pandemic in London. We used an established and validated informatics pipeline to allow rapid evaluation of this important question during the pandemic.

## Methods

This project operated under London South East Research Ethics Committee approval (reference 18/LO/2048) granted to the King’s Electronic Records Research Interface (KERRI); specific work on COVID19 research was reviewed with expert patient input on a virtual committee with Caldicott Guardian oversight.

### Study Design

The study cohort was defined as all adult inpatients testing positive for SARS-Cov2 by RT-PCR at King’s College Hospital and Princess Royal University Hospital from 1st March to 13th April 2020. Only symptomatic patients who required inpatient admission were included. Presenting symptoms included but were not limited to fever, cough, dyspnoea, myalgia, chest pain or delirium. The primary endpoint was defined as death or admission to a critical care unit for organ-support within 21 days of symptoms onset. Data were collected for a range of clinical and demographic parameters (Table 1). To ascertain chronic treatment with ACEi, ARB and other relevant medications, we captured information from clinical notes, outpatient clinic letters and inpatient medication orders. If a drug was a regular medication in the community but withheld on admission, we considered this to be on chronic treatment. The primary endpoint was manually verified by clinician review of the electronic health record.

**Table 1.** Characteristics of the 1200 patients positive for COVID19 at Princess Royal University Hospital and King’s College Hospital, London, UK. All variables were complete and shown as N (% of column) except age which is mean (SD). ACEi = Angiotensin converting enzyme inhibitor; ARB = Angiotensin-2 Receptor Blocker.; HTN = hypertension; HF = heart failure; IHD = ischemic heart disease; COPD = chronic obstructive pulmonary disease; CKD = chronic kidney disease; TIA = transient ischaemic attack; BMI = body mass index. Data was available on all patients except for ethnicity (925), systolic blood pressure (1120), diastolic blood pressure (1120), BMI (621). P-value comparing the group on ACEi /ARB vs. Not On ACEi/ARB with Bonferroni correction for multiple testing. Continuous variables compared with a t-test, binary variables compared with a Chi-squared test.

|  |  | All Patients | On ACEi or<br>ARB | Not on ACEi or<br>ARB | p-value |
| --- | --- | --- | --- | --- | --- |
|  | N | 1200 | 399 | 801 |  |
|  | Age (SD),<br>years | 67.96 (17.07) | 73.02 (13.46) | 65.45 (18.1) | <0.001 |
| Sex | Male | 686 (57.2%) | 231 (57.9%) | 455 (56.8%) | n.s. |
|  | Female | 514 (42.8%) | 168 (42.1%) | 346 (43.2%) | n.s. |
| Ethnicity | Caucasian | 512 (42.7%) | 170 (42.6%) | 342 (42.7%) | n.s. |
|  | Black | 310 (25.8%) | 105 (26.3%) | 205 (25.6%) | n.s. |
|  | Asian | 58 (4.8%) | 21 (5.3%) | 37 (4.6%) | n.s. |
|  | Unknown/<br>Mixed/Other | 320 (26.7%) | 103 (25.8%) | 217 (27.1%) | n.s. |
| Comorbidity | HTN | 645 (53.8%) | 339 (85.0%) | 306 (38.2%) | <0.001 |
|  | Diabetes | 418 (34.8%) | 215 (53.9%) | 203 (25.3%) | <0.001 |
|  | HF | 107 (8.9%) | 65 (16.3%) | 42 (5.2%) | <0.001 |
|  | IHD | 160 (13.3%) | 83 (20.8%) | 77 (9.6%) | <0.001 |
|  | COPD | 121 (10.1%) | 42 (10.5%) | 79 (9.9%) | n.s. |
|  | Asthma | 169 (14.1%) | 58 (14.5%) | 111 (13.9%) | n.s. |
|  | CKD | 206 (17.2%) | 108 (27.1%) | 98 (12.2%) | <0.001 |
|  | Previous Stroke/TIA | 235 (19.6%) | 112 (28.1%) | 123 (15.4%) | <0.001 |
|  | BMI (SD), kg/m<sup>2</sup> | 26.3 (8.7) | 27.0 (8.5) | 25.8 (8.4) | n.s. |
|  | BMI≥30 | 182 (15.2%) | 69 (17.3%) | 113 (14.1%) | n.s. |
| Number of comorbidities | 0 | 310 (25.8%) | 19 (4.8%) | 291 (36.3%) | <0.001 |
|  | 1 | 283 (23.6%) | 73 (18.3%) | 210 (26.2%) | 0.08 |
|  | >1 | 607 (50.6%) | 307 (76.9%) | 300 (37.5%) | <0.001 |
| Drugs | ACEi | 260 (21.7%) | 260 (65.2%) | 0 (0.0%) |  |
|  | ARB | 147 (12.2%) | 147 (36.8%) | 0 (0.0%) |  |
|  | Statin | 472 (39.3%) | 268 (67.2%) | 204 (25.5%) | <0.001 |
|  | Beta-blocker | 337 (28.1%) | 184 (46.1%) | 153 (19.1%) | <0.001 |
| Vital Signs | Systolic BP (SD), mmHg | 124 (27) | 126 (28) | 123 (26) | 0.17 |
|  | Diastolic BP (SD), mmHg | 71 (18) | 71 (18) | 71 (18) | n.s. |
| Primary endpoint by 21 days | Death or Critical Care admission | 415 (34.6%) | 127 (31.8%) | 288 (36.0%) | n.s. |
|  | Death | 288 (24.0%) | 106 (26.6%) | 182 (22.7%) | n.s. |
|  | Critical Care admission and<br>Alive | 127 (10.6%) | 21 (5.3%) | 106 (13.2%) | <0.01 |

### Data Processing

The data (demographic, emergency department letters, discharge summaries, clinical notes, radiology reports, medication orders, lab results) was retrieved and analysed in near real-time from the structured and unstructured components of the electronic health record (EHR) using a variety of well-validated natural language processing (NLP) informatics tools belonging to the CogStack ecosystem,^11^ namely DrugPipeline,^12^ MedCAT^13^ and MedCATTrainer.^14^ The CogStack NLP pipeline captures negation, synonyms, and acronyms for medical SNOMED-CT concepts as well as surrounding linguistic context using deep learning and long short-term memory networks. DrugPipeline was used to annotate medications and MedCAT produced unsupervised annotations for all SNOMED-CT concepts under parent terms Clinical Finding, Disorder, Organism, and Event with disambiguation, pre-trained on MIMIC-III.^15^ Further supervised training improved detection of annotations and meta-annotations such as experiencer (is the concept annotated experienced by the patient or other), negation (is the concept annotated negated or not) and temporality (is the concept annotated in the past or present) with MedCATTrainer. Meta-annotations for hypothetical and experiencer were merged into Irrelevant meaning that any concept annotated as either hypothetical or where the experiencer was not the patient was annotated as irrelevant. Performance of the MedCAT NLP pipeline for disorders mentioned in the text was evaluated on 5617 annotations for 265 documents by a domain expert (JTHT) and F1, precision and recall recorded. Additional full case review for correct subsequent diagnosis assignment was performed by 3 clinicians (JTHT, KOG, RZ) for key comorbidities. The performance of DrugPipeline has previously been described.^12^ Manual review of 100 detections gave F1=0.91 for exclusion of drug allergies by DrugPipeline.

### Statistical Analysis

In order to investigate the association between ACEi/ARB and disease severity measured as critical care admission or death, we performed a series of logistic regressions. In a first step, we explored independently the association for ACEi/ARB (Baseline model). In a second step, we adjusted the model for age and sex (Model 1). Then, we additionally adjusted for hypertension (Model 2) and finally, additionally adjusted for other comorbidities, i.e. diabetes, ischemic heart disease, heart failure and chronic kidney disease (Model 3). We also explored the independent association for hypertension following the same modelling approach. In addition, we assessed the robustness to unmeasured confounders of the fully adjusted estimate of ACEi/ARB effect using the e-value approach, which are defined as the minimum strength of association on the risk-ratio scale that an unmeasured confounder would need to have with both the treatment assignment and the outcome to fully explain away a specific treatment-outcome association, conditional on the measured covariates^16^. Sensitivity analyses were performed i) requiring at least two detections of medication for positive exposure; ii) using only structured data on in-hospital medication orders; iii) ignoring our 21 day window for medications; iv) testing sensitivity to unmeasured confounders.

## Results

Our total cohort consisted of 1200 confirmed positive symptomatic inpatients aged 63+20 (SD) years with 52% being male (**Table 1**). The patients were of diverse ethnicities with over 30% from minority ethnic groups. Nearly 75% of patients had one or more comorbidities. The commonest comorbidities were hypertension (51.2%), diabetes (30.2%), chronic kidney disease (17.2%), and ischaemic heart disease or heart failure (22.2%). 15.2% of patients had a BMI greater than 30 kg/m^2^. A total of 399 patients (33.2%) were on chronic treatment with ACEi or ARB. 415 of the 1200 patients (34.6%) required admission to the Critical Care Unit or had died within 21 days of symptom onset. Among patients who achieved the primary endpoint (Death or Critical Care admission), the percentage who had positive mentions for various disorders derived via the NLP for medical concept annotations with F1 > 80% and more than 10 annotated mentions, as compared to those not achieving the primary endpoint, is shown in Figure 1. The performance of the NLP pipeline is shown in Figure 2. Manual validation of the presence of comorbidities was performed in a sample of 200 patients and showed excellent performance, for example a false positive rate of 1% for hypertension and 0% for diabetes.

**Figure 1.**
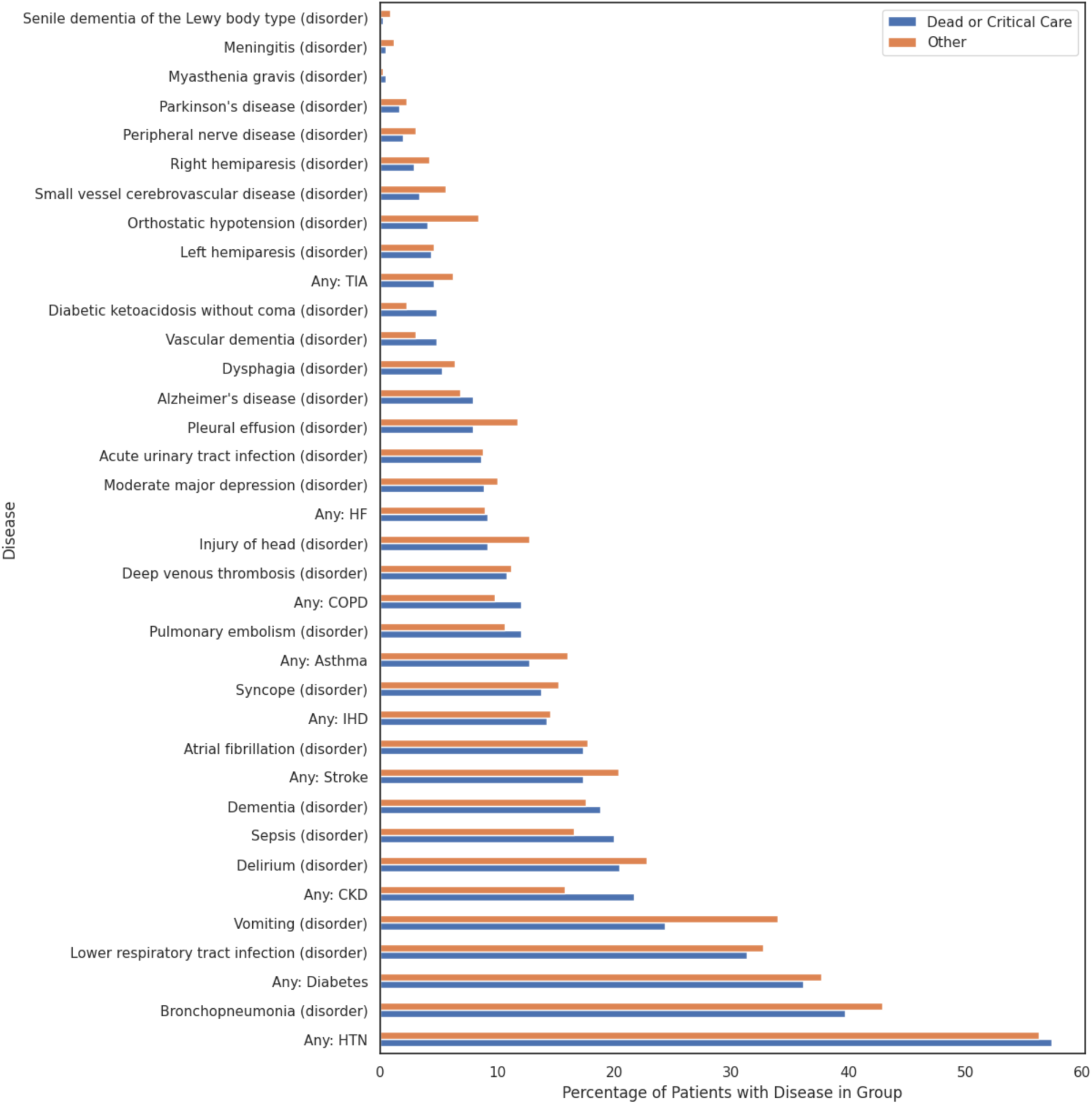
Distribution of disorders between patients achieving the primary outcome (Death or Critical Care admission) and those not achieving it by 21 days after symptom onset. The percentage of patients that have a positive mention of a disorder in each of the two groups is shown. All diseases were extracted from free-text using Cogstack and MedCAT. Only medical concept annotations with F1 > 80% and more than 10 annotated samples are shown. Disease names that start “Any:” are aggregate concepts for multiple specific conditions that are used in our analysis.

**Figure 2.**
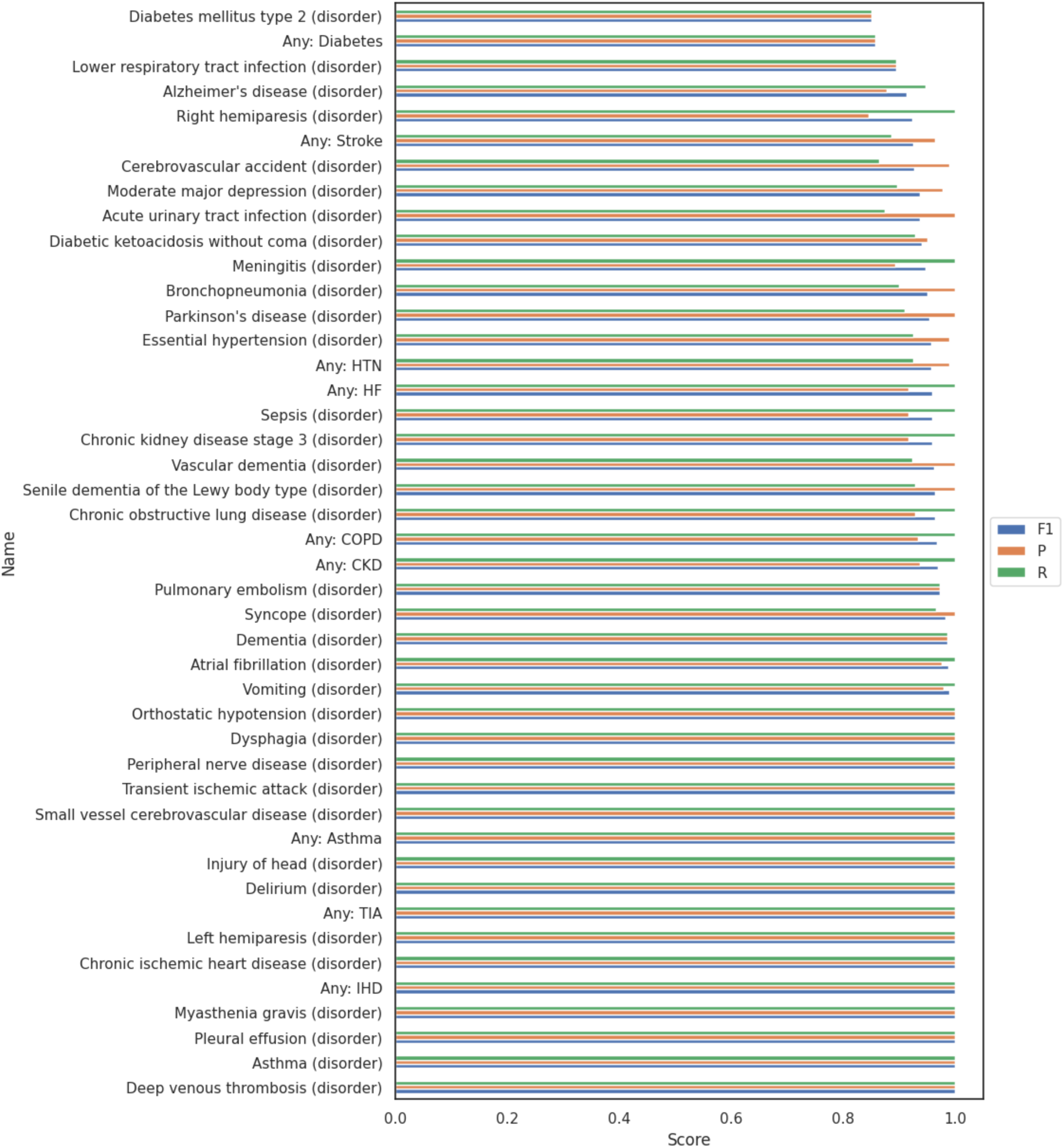
Performance of the CogStack and MedCAT NLP pipeline in detecting disease mentions within the electronic health record text. Precision (P), Recall (R) and F1 (harmonic mean of precision and recall). Only medical concept annotations with F1 > 80% and more than 10 annotated samples are shown. Disease names that start “Any:” are aggregate concepts for multiple specific conditions that are used in our analysis.

We next compared the outcome of patients on chronic treatment with ACEi/ARB versus those not on these agents. The group on ACEi/ARB were significantly older but had a similar male/female split and a similar ethnicity profile to those not on ACEi/ARB (**Table 1**). The BMI was similar between groups. There was a greater proportion of patients with cardiovascular comorbidities (hypertension, diabetes, heart failure, ischaemic heart disease) and chronic kidney disease in the group on ACEi/ARB than those not taking these drugs, as would be expected. Therefore, the patients on ACEi/ARB had a higher prevalence of factors associated with worse outcome of COVID19 disease in prior studies.^1–3^ The ACEi/ARB group also had higher rates of treatment with beta blockers and statins than those not on ACEi/ARB, consistent with their higher rates of cardiovascular morbidities. **Figure 3** shows Kaplan-Meier curves for the primary end-point in patients on ACEi/ARB and those not on these drugs.

**Figure 3.**
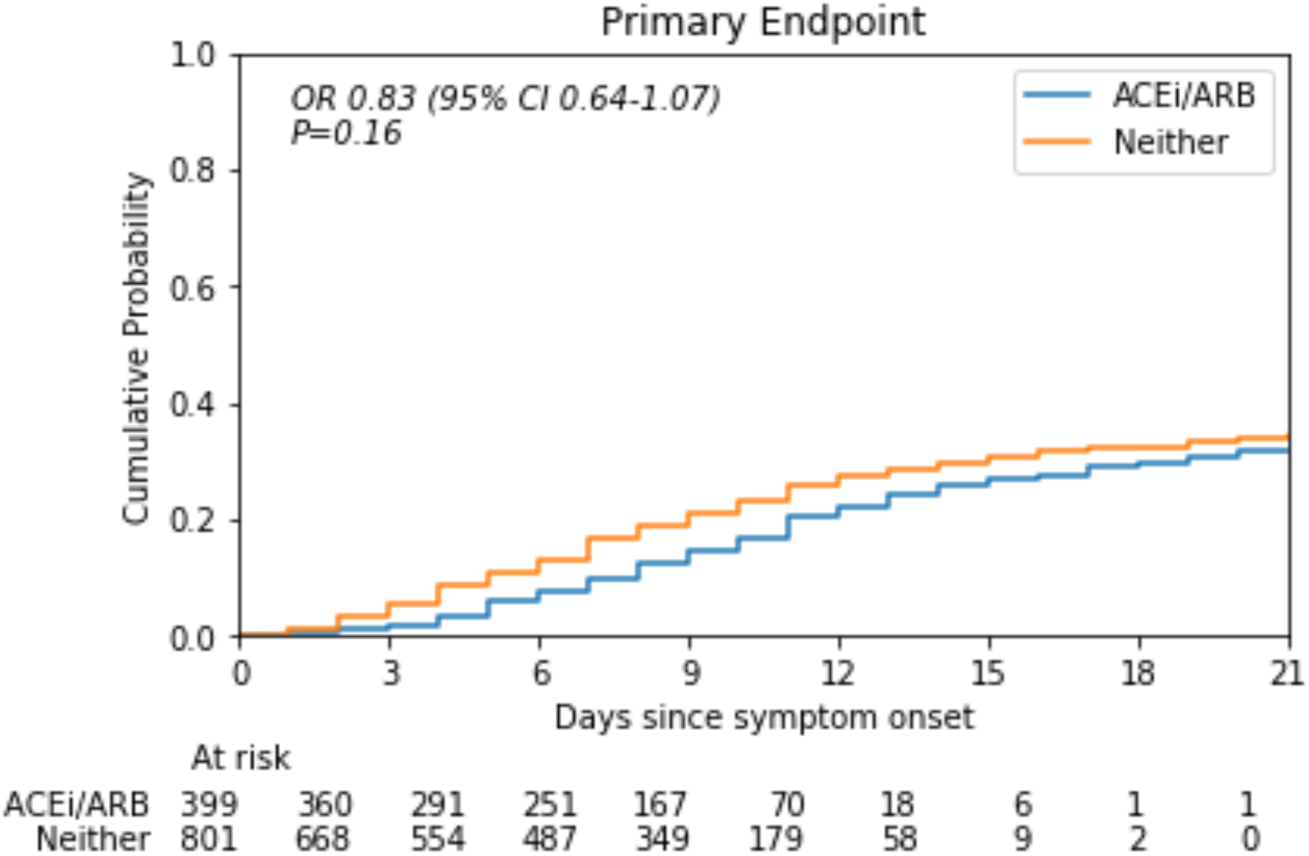
Kaplan-Meier Curves for the primary endpoint in COVID19 patients on chronic treatment with ACE-inhibitors (ACEi) or Angiotensin-II Receptor Blockers (ARB) (blue line) versus those not on these drugs (orange line) The unadjusted Odds Ratio (OR) for the primary endpoint for those on ACEi/ARB was 0.83 (p =0.16); the adjusted OR was 0.63 (p<0.01).

To assess the independent effect of ACEi/ARB on primary outcome, we first performed an unadjusted logistic regression analysis. This indicated that the likelihood of a severe outcome was similar in individuals on ACEi/ARB as compared to those not on these drugs, with an Odds Ratio (OR) of 0 83 (CI 0-64-1·07) - Baseline Model, **Table 2**. However, after adjustments for age and sex (Model 1 in **Table 2**), the likelihood of severe disease was significantly lower in those on ACEi/ARBs (OR 0.70 [0.53-0.91], p<0.01). Additional adjustment for hypertension (Model 2 in **Table 2**) and for the other major comorbidities, diabetes, chronic kidney disease, and ischaemic heart disease/heart failure (Model 3 in **Table 2**), had a modest further effect. The OR for the primary outcome in Model 3 was 0.63 (0.47-0.84), p<0.01. **Supplementary Table 1** shows the OR and p-values for all variables in each model. Male patients were found to have a higher likelihood of severe disease in Model 3 (OR 1·50 [CI 1·17-1·93], p=<0.01).

**Table 2.** Summary of Odds Ratios for ACE inhibitor and/or ARB drug use and the primary endpoint. Odds ratios (OR) and p-values calculated from logistic regressions. ACEi = Angiotensin converting enzyme inhibitor; ARB = Angiotensin-2 Receptor Blocker.

| Model | Adjustments | OR (95% CI)<br>ACEi/ARB<br>vs neither drug | p |
| --- | --- | --- | --- |
| Baseline | - | 0.83 (0.64-1.07) | 0.16 |
| Model 1 | Age, sex | 0.70 (0.53-0.91) | <0.01 |
| Model 2 | Age, sex, hypertension | 0.64 (0.48-0.86) | <0.01 |
| Model 3 | Age, sex, hypertension, diabetes mellitus, chronic kidney disease, ischaemic heart disease, heart failure | 0.63 (0.47-0.84) | <0.01 |

We also examined the independent association between hypertension and disease severity. The results showed that individuals with hypertension had a similar likelihood of suffering a severe outcome as those that without hypertension, either in unadjusted models (OR 1·25 [CI 0·98-1·59]; p=0·069) or in models adjusted for age and gender (OR 1·03 [CI 0·80-1·32]; p=0·83).

Sensitivity analyses were performed using criteria for ACEi exposure that were either more strict (requiring multiple mentions in the clinical notes or using only in-hospital medication orders as evidence) or less strict (including any mention of ACEi treatment even outside a 21 day window from onset of symptoms). In all cases, the estimates of the impact of ACEi treatment were consistent with those in **Table 2**. In analysis requiring at least two mentions of chronic treatment with ACEi/ARB, we found that this was significant in the uncorrected baseline model (a lower OR). We estimated an e-value of 1.818 which suggests that the estimate could be vulnerable to possible confounders not yet included.

## Discussion

This study in a large consecutive cohort of 1200 patients in the UK suggests that chronic treatment with ACEi and ARB is not associated with an increase in severe outcome of COVID19 disease, defined as death or admission to a critical care unit. The hypothetical relationship between treatment with ACEi/ARB and severe COVID19 disease has been intensely debated.^6-7,8^ There are theoretical mechanisms whereby chronic treatment with ACEi/ARB might increase propensity to SARS-CoV2 infection as well as other mechanisms whereby treatment with these agents might be beneficial. It is a particularly important question because chronic treatment with ACEi/ARB is of proven benefit in conditions such as hypertension, diabetes, chronic kidney disease and heart failure and an unwarranted cessation of therapy in patients with these conditions as a result of the SARS-CoV2 pandemic could have serious long-term detrimental effects.

The general clinical characteristics and the rates of severe outcome of the patients in our study were broadly similar to those that have been described in recent large series from Italy and the USA.^1–3^ We found that patients who were on chronic treatment with ACEi/ARB had many demographic and comorbidity features that have been associated in previous studies with worse outcome in COVID-19 disease, such as an older age and a higher prevalence of hypertension, diabetes, heart failure and other morbidities ^1–3^. Treatment with ACEi/ARB was nevertheless not associated with an increase in rates of severe outcomes, with or without adjustment for age, sex and comorbidities. Two very recent studies from China have reported on the relationship between ACEi/ARB and outcome of COVID-19 disease in hospitalised patients. In a single centre study from Wuhan in which only 115 of 1178 patients (<10%) were taking ACEi/ARB, the authors did not find any relationship between these drugs and outcome,^17^ the data are however limited by the low numbers on ACEi/ARB and potential confounding by other factors. A second report was a retrospective multi-centre study including 1128 patients but again had only 188 patients (16.6%) on treatment with ACEi/ARB.^18^ This study suggested that treatment with ACEi/ARB was associated with a lower rate of severe outcome with COVID19 infection. Our study is the first to be conducted on an ethnically mixed population in the western world and includes significant proportions of both White and minority ethnic (Black, Asian) patients. The rates of usage of ACEi/ARB in our study (33.2%) are in line with those expected in well-treated patients with comorbidities and are therefore, in principle, more applicable to patients in Europe and the Americas. Ethnicity is a very pertinent issue in this regard due to the recognised ethnicity-related differences in response to drugs affecting the RAS.^19,20^ Of relevance, the ethnicity profiles of the patients on ACEi/ARB in our study were similar to those not taking these drugs.

In the current study, when we adjusted for age, sex and comorbidities in logistic regression analyses, the OR for a severe outcome was significantly lower in patients on ACEi/ARB than those not on these agents. This suggestion of a favourable association of treatment with ACEi/ARB and less severe outcome in COVID19 disease would be consistent with the hypothesised beneficial effects of inhibition of RAS activation in patients with severe lung injury or Acute Respiratory Distress Syndrome (ARDS).^7,8^ However due to possibility of unmeasured confounds, the confirmation of a potential therapeutic benefit of treatment with ACEi/ARB in COVID19 disease would require further studies and randomised control trials.

This study used an NLP approach to perform very rapid analysis of high volume, unstructured real world clinical data. This however introduces the possibility of missing circumlocutory mentions of disease, symptoms or medications. We mitigated against this by manually validating annotations in a subset of records and also verifying drug treatments against inpatient electronic prescription data. Moreover, we performed sensitivity analyses to test the impact of different criteria to define the ACEi/ARB exposed cohort on our results, and found that the OR remained <1.0 and significant for ACEi/ARB exposure in all adjusted analyses. We therefore consider our analysis pipeline to be robust to specific details of the pipeline that are not clinically relevant. However we did find that the estimated odds ratio may be sensitive to unmeasured confounding, which suggests caution in the interpretation of any protective effect and the need for replication in a larger sample remains.

Our study has some potential limitations. Although the patients and data were prospectively collected, the analyses were retrospective. The study was conducted on two hospital sites in a single geographical, albeit ethnically mixed, locus in the UK over a relatively short follow-up period. However, the duration of follow-up is sufficient to accurately detect early severe outcomes based on the data from multiple studies during the current pandemic. We used the covariates identified as important in the previous large case series on COVID19 ^1–3^, including age, sex and common comorbidities, to adjust our analyses. However, it is possible that other unmeasured confounders could have influenced the results. For example, the patients on chronic ACEi/ARB treatment were also more frequently treated with statins than those not on these drugs, which could suggest that their medical conditions were generally better managed. However, the ACEi/ARB group was also older and had higher rates of hypertension, diabetes and multiple morbidities, making it unlikely that these patients were physiologically healthier. Our study was performed in patients with COVID19 who required hospitalisation; the effect of chronic treatment with ACEi/ARB on less severe infection with SARS-CoV2 in the non-hospital setting requires further study. Whether the current results are applicable to other global populations, such as in Africa, will also require additional study.

In summary, the results of this study in 1200 patients show no evidence of a detrimental effect of chronic treatment with ACEi/ARB in patients presenting with severe COVID19 infection. Patients on treatment with ACEi/ARB should continue these drugs during their COVID19 illness as per current ESC Council guidelines.^21^

## Data Availability

Source text from
patient records used in the study will not be
available due to inability to fully anonymise up to
the Information Commissioner Office (ICO)
standards and would be likely to contain strong
identifiers (e.g. names, postcodes) and highly
sensitive data (e.g. diagnoses). A subset of the
dataset limited to anonymisable information (e.g.
only UMLS codes and demographics) is available
on request to researchers with suitable training in
information governance and human confidentiality
protocols subject to approval by the King’s College
Hospital Information Governance committee;

## Acknowledgements

DMB is funded by a UKRI Innovation Fellowship (Health Data Research UK MR/S00310X/1). RB is funded in part by the King’s College London UK Medical Research Council (MRC) Skills Development Fellowship programme (MR/R016372/1) and the National Institute for Health Research (NIHR) Biomedical Research Centre at South London and Maudsley NHS Foundation Trust and King’s College London (SLAM-KCL; IS-BRC-1215-20018). AF is supported by the NIHR Biomedical Research Centre at SLAM-KCL (IS-BRC-1215-20018) and the NIHR centre at University College London Hospitals. RJBD is supported by Health Data Research UK; the BigData@Heart Consortium, funded by the Innovative Medicines Initiative-2 Joint Undertaking (Grant No. 116074) under the European Union Horizon 2020 programme; the NIHR University College London Hospitals Biomedical Research Centre; the NIHR Biomedical Research Centre at SLAM-KCL. KO’G is supported by an MRC Clinical Training Fellowship. RZ is supported by a King’s Prize Fellowship. AS is supported by a King’s Medical Research Trust studentship. AMS is supported by the British Heart Foundation (CH/1999001/11735), the NIHR Biomedical Research Centre at Guy’s & St Thomas’ NHS Foundation Trust and King’s College London (IS-BRC-1215-20006), and the Fondation Leducq. AP is partially supported by NIHR NF-SI-0617-10120. This work was supported by the NIHR University College London Hospitals Biomedical Research Centre Clinical and Research Informatics Unit; NIHR Health Informatics Collaborative; the Institute of Health Informatics at University College London; and Health Data Research UK. The manuscript represents independent research part-funded by the NIHR Biomedical Research Centre at SLAM-KCL. The views expressed are those of the authors and not necessarily those of the NHS, the NIHR or the Department of Health and Social Care. The funders had no role in study design, data collection and analysis, decision to publish, or preparation of the manuscript. We thank all the clinicians managing the patients, the patient experts of the KERRI committee, AI4VBH, Professor Irene Higginson, Professor Alastair Baker, Professor Jules Wendon, Dan Persson and Damian Lewsley for their support.

## Author Contributions

JTHT, RJBD, DMB, ZK, TS, AF conceived the study design

DMB, ZK, AS, TS, JTHT, LR, KN performed data processing and software development

KOG, RZ, JTHT performed data validation

DMB, AP, RB performed statistical analysis

JTHT, DMB, ZK, TS, AF, RJBD, AMS performed critical review

JTHT, RJBD, DMB, AMS, ZK, TS, AF, DMB, AP, RB wrote the manuscript

## Supplementary files

**Supplementary Table 1.** Odds ratios and p-values for all variables and primary endpoint. Odds ratios and p-values calculated from logistic regressions. ACEi = Angiotensin converting enzyme inhibitor. OR = Odds ratio. ARB = Angiotensin Receptor Blocker. HTN = hypertension; HF = heart failure; IHD = ischaemic heart disease; CKD = chronic kidney disease.

| Model | Variable | OR (95% CI) | P-value |
| --- | --- | --- | --- |
| Baseline | On ACEi or ARB | 0.83 (0.64-1.07) | 0.16 |
| Model 1 | On ACEi or ARB | 0.70 (0.53-0.91) | <0.01 |
|  | Age (per 10 years) | 1.26 (1.17-1.36) | <0.01 |
|  | Male | 1.51 (1.18-1.93) | <0.01 |
| Model 2 | On ACEi or ARB | 0.64 (0.48-0.86) | <0.01 |
|  | Age (per 10 years) | 1.25 (1.16-1.35) | <0.01 |
|  | Male | 1.51 (1.18-1.94) | <0.01 |
|  | HTN | 1.22 (0.92-1.60) | 0.16 |
| Model 3 | On ACEi or ARB | 0.63 (0.47-0.84) | <0.01 |
|  | Age (per 10 years) | 1.24 (1.14-1.34) | <0.01 |
|  | Male | 1.50 (1.17-1.93) | <0.01 |
|  | HTN | 1.15 (0.86-1.55) | 0.34 |
|  | Diabetes | 1.07 (0.81-1.40) | 0.64 |
|  | HF or IHD | 0.95 (0.68-1.31) | 0.73 |
|  | CKD | 1.33 (0.95-1.86) | 0.09 |
| Hypertension unadjusted | HTN | 1.25 (0.98-1.59) | 0.069 |
| Hypertension adjusted | Age (per 10 years) | 1.24 (1.15-1.33) | <0.01 |
|  | Male | 1.50 (1.17-1.92) | <0.01 |
|  | HTN | 1.03 (0.80-1.32) | 0.83 |

